# Evidence that Telomere Length is Causal for Idiopathic Pulmonary Fibrosis but not Chronic Obstructive Pulmonary Disease: A Mendelian Randomisation Study

**DOI:** 10.1101/2020.02.05.20019653

**Authors:** Anna Duckworth, Michael A. Gibbons, Richard J. Allen, Howard Almond, Robin N. Beaumont, Andrew R. Wood, Katie Lunnon, Mark A. Lindsay, Louise V. Wain, Jess Tyrrell, Chris J. Scotton

## Abstract

**Background:** Idiopathic pulmonary fibrosis (IPF) is a fatal lung disease accounting for 1% of UK deaths. In the familial form of pulmonary fibrosis, causal genes have been identified in ∼30% of cases, and a majority relate to telomere maintenance. Prematurely shortened leukocyte telomere length has also been associated with IPF, as well as chronic obstructive pulmonary disease (COPD), a disease with a similar demographic and shared risk factors. Using Mendelian randomisation (MR), our study aimed to determine whether short telomeres cause IPF or COPD.

**Methods:** We performed an MR study for telomere length causality in IPF and COPD with up to 1,369 IPF cases, 14,103 COPD cases and 435,866 controls of European ancestry in UK Biobank. Initial studies using polygenic risk scores followed by two-sample MR analyses were carried out using seven genetic variants previously associated with telomere length, with replication analysis in an IPF cohort of 2,668 IPF cases and 8,591 controls and a COPD cohort of 15,256 cases and 47,936 controls.

**Findings:** Meta-analysis of the two-sample MR results provided evidence that shorter telomeres cause IPF, with a genetically instrumented one standard deviation shorter telomere length associated with 5.81 higher odds of IPF ([95% CI: 3.56-9.50], P=2.19×10^−12^. Despite being an age-related lung disease with overlapping risk, there was no evidence that telomere length caused COPD (OR 1.07, [95% CI 0.90-1.27], P = 0.46).

**Interpretation:** Cellular senescence is hypothesised as a major driving force in both IPF and COPD; telomere shortening may be a contributory factor in IPF, suggesting divergent mechanisms in COPD. This enables greater focus in telomere-related diagnostics, treatments and the search for a cure in IPF. Therapies manifesting improvements in telomere length, including safe telomere activation therapy, may warrant investigation.

## Introduction

Idiopathic pulmonary fibrosis (IPF) is a complex and incurable fibrotic lung disease. Based on data from 2012 [1], the average lifespan following diagnosis is widely quoted as being around 3yrs and IPF accounts for around 5300 UK deaths each year, over 1% of all UK deaths.

### The association of PF and COPD with telomere length

In approximately 20% of cases, pulmonary fibrosis (PF) clusters in family groups [2]. Inherited genetic causes have been established for around 30% of familial cases and the majority relate to telomere maintenance. The most common of these is the gene encoding telomerase reverse transcriptase (TERT), a catalytic subunit of the enzyme telomerase which works in conjunction with the telomerase RNA component, TERC [3]. Variants in these two genes account for 19% of familial cases via autosomal dominant inheritance with reduced penetrance.

Idiopathic pulmonary fibrosis risk also has a strong genetic component [4]. Prematurely shortened leukocyte telomere length (LTL) has been associated with IPF and also chronic obstructive pulmonary disease (COPD), a condition with a similar demographic. Studies have shown age adjusted LTL values of 0.85 ± 0.60 vs 1.15 ± 0.6, p=0.0001 relative to reference DNA for IPF versus controls [5] and 0.68 ± 0.25 vs. 0.88 ± 0.52, p = 0.003 for COPD versus smoking controls [6]. There is also evidence for an association between shorter leukocyte telomere length and worsened survival in IPF [7], with the suggestion that LTL could be used as a predictive biomarker. A recent study which included 32 IPF patients undergoing diagnostic lung biopsy demonstrated shortened lung telomeres (particularly in type II alveolar epithelial cells within fibrotic lesions), but no correlation with age; half of these showed excessive lung telomere shortening, to the same extent seen in PF patients with a TERT mutation – suggestive of a disease driven by telomere attrition [8].

The most widely used model for experimental exploration and pre-clinical assessment for IPF is the murine model of bleomycin-induced lung injury[9]. Although this mouse model has been an important precursor to clinical trials and has helped enable the development and licensing of the two main IPF treatments, Nintedanib and Pirfenidone, the majority of studies utilise young mice where the fibrotic lesions lack features which are characteristic of IPF (such as honeycombing and fibrotic foci), and where the fibrosis may resolve depending on the severity of initial injury [10]. Mice have particularly long telomeres; studies which have utilised aged mice (>18mo old) have demonstrated a more profound fibrotic response which does not resolve [11], and which better reflects the human disease. Notably, a recent study of low-dose bleomycin in *TERT*^*-/-*^ mice (which have significantly shortened telomeres) demonstrated that targeted telomerase activation in type II alveolar epithelial cells (AEC2s) cells using gene therapy with adeno-associated vectors (AAV) showed therapeutic effects in mice with established fibrosis [12] through telomere elongation and increased proliferation of AEC2 cells combined with lower DNA damage, apoptosis and senescence burden.

Thus, while there is evidence of an association with shorter telomeres in both diseases, experimentally, there is currently only evidence from murine models of PF that telomere extension is therapeutic.

### Mendelian Randomisation

A genetic technique known as Mendelian randomisation (MR) [13] can be used to test for a causal relationship between a phenotype that can be genetically influenced (such as telomere length) and a disease outcome, such as IPF or COPD. Causality in one direction is shown because genetic make-up is allocated at conception and unlikely to be influenced by disease in later life. Potential confounding influences such as smoking, pollution and other environmental/lifestyle risk factors are removed from the analysis, creating in effect a natural blind randomised control trial (Figure 1). Recent genome-wide association studies (GWAS) [14, 15] have identified several genetic variants or single nucleotide polymorphisms (SNPs) that are independently associated with telomere length and provide potential tools for MR. A previous study investigating associations between genetically increased telomere length and risk of cancer and other non-neoplastic diseases, reported increased risk of site-specific cancers and reduced risk of coronary artery disease, celiac disease and interstitial lung diseases [16]. We therefore hypothesised that telomere length is causally linked to IPF but not COPD, given that inherited genetic defects in telomerase production lead to familial pulmonary fibrosis. To test this, we used MR and the latest data release from up to 451,025 participants in the UK Biobank together with a genetic instrument associated with shorter telomere length. We further tested the effect in males and females separately since there is a well-established gender bias in IPF.

**Figure 1.**
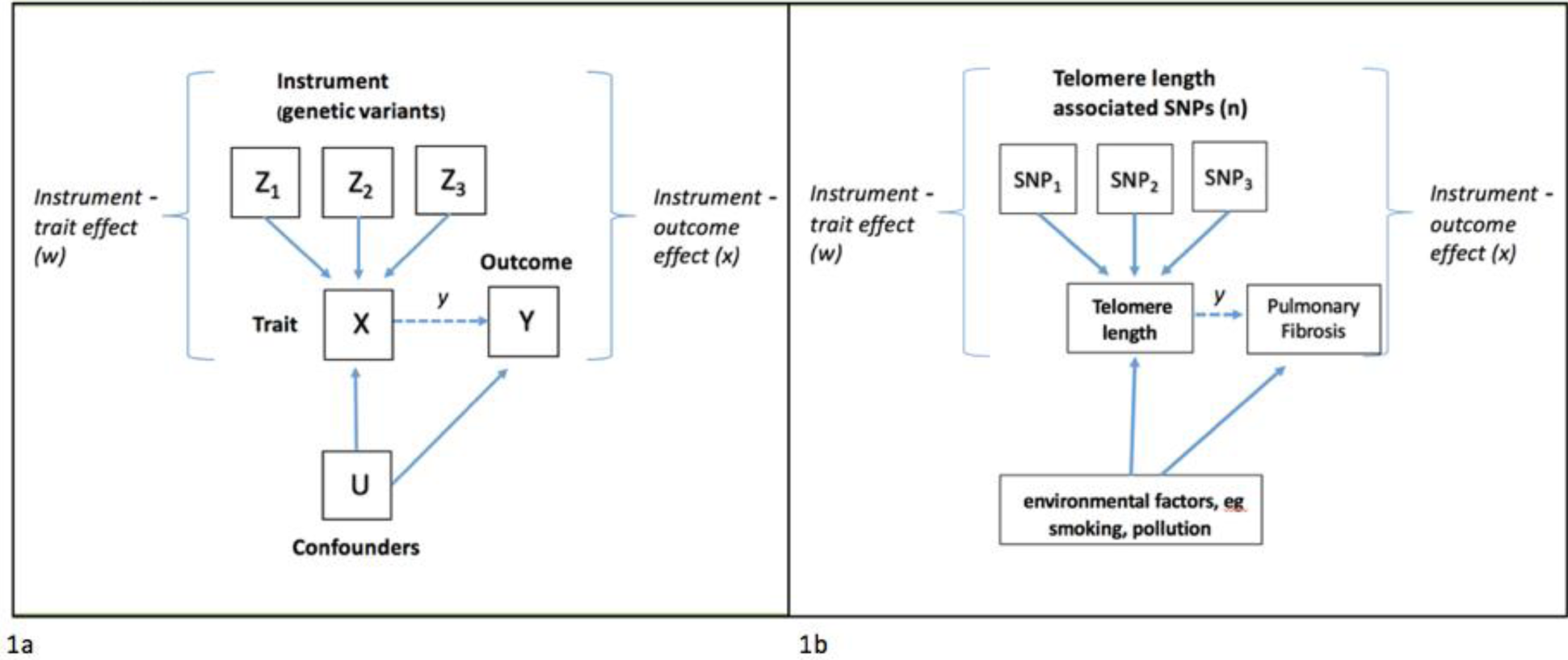
Principle of Mendelian randomisation; if telomere length plays a causal role in IPF, genetic variants associated with telomere length will also be associated with IPF. 1a) If the observed trait X causes the particular outcome Y, the instrument Z (genetic variants associated with the trait) will also be associated with the outcome. In our case, if telomere length causes idiopathic pulmonary fibrosis, genetic variants associated with telomere length will also be associated with IPF. As genotype is assigned at conception, it should not be associated with risk factors that might confound the association between telomere length and IPF (eg smoking or pollution). We can use weighted estimates of the genetic-telomere length association (w) and the genetic IPF-association (x) to infer the causal effect of telomere length on IPF (y-x/w), which is expected to be free from confounding.

## Methods

### Collection and selection of UK Biobank Data

The UK Biobank is a study of 500,000 volunteers aged between 37-73 years, recruited across the UK during 2006-2010 [17], with participant data including physical measurements, biological samples (blood, urine and saliva) for biomarker and genetic analysis, and long-term follow-up via hospital record linkage. Information on patient and public involvement is available online [18]. The cut-off date for data in this study was 31/3/2017. Genetic variant or single nucleotide polymorphism (SNP) data was generated from the Affymetrix Axiom UK Biobank array (for ∼450,000 individuals) and the UK BiLEVE array (∼50,000 individuals) following extensive quality control, as described previously [19]. We identified two UK Biobank study populations: 1) The full set of available participants (451,025 individuals of European ancestry), including related individuals, to maximise statistical power in the 2-sample Mendelian randomisation where relatedness is handled within the model; 2) A smaller subset of 379,708 unrelated individuals (defined using a KING Kinship, generating an optimal list of unrelated individuals with maximum inclusion), important for the initial regression modelling. Ancestral principal components were then generated within these identified individuals for use in subsequent analyses, as described previously [20].

IPF cases within UK Biobank were defined as those having a primary or secondary ICD10 code HES (Hospital Episodes Statistics) diagnosis of J84.1. With this ‘narrow’ criterion, we identified 1369 cases (1133 unrelated, whereby no two participants are third degree related or closer). We repeated our analysis with a ‘broad’ IPF definition that included J84.0 (Alveolar and parieto-alveolar conditions), J84.8 (Other specified interstitial pulmonary diseases) or J84.9 (Interstitial pulmonary disease, unspecified) and using these criteria, we identified 1,621 (1,355 unrelated) cases. COPD cases were defined as those having a primary or secondary ICD10 code of J41 (Simple and mucopurulent chronic bronchitis), J42 (Unspecified chronic bronchitis), J43 (Emphysema) or J44 (Other chronic obstructive pulmonary disease) plus those self-reported to have COPD: a total of 14,103 (11,895 unrelated) cases. Using the broad definition of IPF, we subtracted 566 (482 unrelated) cases with both IPF and COPD to leave 13,538 (11,413 unrelated) cases.

For the control group, we removed both COPD cases and broad definition IPF cases to give 435,866 (366,942 unrelated) clean controls.

### Replication cohorts

An IPF replication cohort was derived from the discovery stage of a recent GWAS study which comprised three independent case-control studies from the UK, Chicago and Colorado a total of 2,668 IPF cases and 8,591 controls [21]. The UK study included matched controls selected from UK Biobank.

The COPD replication cohort was derived from a recent COPD GWAS study with 15,256 cases and 47,936 controls, with 94% of cases and 83% of controls being of European ancestry [22].

### Observational Associations

We used logistic regression models in Stata 13.0 to compare the key demographics of the IPF and COPD groups with controls, adjusting for age and sex as appropriate. These analyses were performed in the unrelated subset of individuals to prevent familial bias.

### Identification of genetic instrument variants

Genetic variants for telomere length were chosen from published GWAS studies, using GWAS that did not include data from the UK Biobank. We used an instrument composed of 7 variants robustly associated with leukocyte telomere length derived from a genome-wide meta-analysis of 37,684 individuals of European descent, with replication of selected variants in a further 10,739 individuals by Codd *et al* [14] (see Supplementary Material, Table 1a). Other telomere length GWAS results were available (see [16]) but with smaller sample sizes and several of the variants available were in linkage disequilibrium (Supplementary Data, Table 1b) so did not meet our MR criteria.

**Table 1a:** Demographics for Idiopathic Pulmonary Fibrosis cases using narrow J84.1 ICD10 code definition, from unrelated individuals of European ancestry in UK Biobank derived using logistic regression analyses. (Odds ratios and p values are adjusted for age and sex, other than percent predicted spirometry which already includes adjustment).

| Demographic | IPF J84.1 only | Controls | OR | 95% CI |  | P |
| --- | --- | --- | --- | --- | --- | --- |
| N = 368,075 | 1,133 | 366,942 |  |  |  |  |
| Mean age at baseline (SD) | 63.1 (5.8) | 57.1 (8.0) | 1.13 | 1.12 | 1.14 | <1X10<sup>-15</sup> |
| Mean age at diagnosis (SD) | 67.2 (7.5) |  |  |  |  |  |
| Male sex, N (%) | 718 (63.4%) | 167,910 (45.8%) | 1.94 | 1.71 | 2.19 | <1X10<sup>-15</sup> |
| Townsend Deprivation Index (SD) | -0.80 (3.33) | -1.53 (2.95) | 1.10 | 1.08 | 1.12 | <1X10<sup>-15</sup> |
| Pollution (SD) | 27.2 (8.0) | 26.2 (7.4) | 1.02 | 1.02 | 1.03 | 6.0x10<sup>-10</sup> |
| Smoking status |  |  |  |  |  |  |
| Never smoker | 341 (30.1%) | 201,987 (55.1%) |  |  |  |  |
| Former smoker | 602 (53.1%) | 128,242 (35.0%) | 2.14 | 1.87 | 2.45 | <1X10<sup>-15</sup> |
| Current smoker | 167 (14.7%) | 32,022 (8.73%) | 3.42 | 2.83 | 4.13 | <1X10<sup>-15</sup> |
| Missing | 23 (2.0%) | 5,104 (1.3%) |  |  |  |  |
| Income (SD) | 1.97 (1.03) | 2.66 (1.19) | 0.69 | 0.65 | 0.74 | <1X10<sup>-15</sup> |
| Mean FEV1 (SD) | 2.36 (0.69) | 2.77 (0.77) | 0.35 | 0.32 | 0.40 | <1X10<sup>-15</sup> |
| FEV1 percent predicted (SD) | 85.0 (23.5) | 91.1 (22.6) | 0.99 | 0.98 | 0.99 | <1X10<sup>-15</sup> |
| Mean FVC (SD) | 3.18 (0.88) | 3.66 (1.00) | 0.42 | 0.38 | 0.46 | <1X10<sup>-15</sup> |
| FVC percent predicted (SD) | 114.8(29.6) | 120.2 (29.6) | 0.99 | 0.99 | 1.00 | <1X10<sup>-15</sup> |
| Physical activity score (SD) | 7.07 (1.31) | 7.41 (1.13) | 0.76 | 0.73 | 0.81 | <1X10<sup>-15</sup> |
| *Participants deceased, N (%)* | *439 (38.8%)* | *10,977 (3.0%)* | *11.8* | *10.4* | *13.4* | *<1X10<sup>-15</sup>* |

**Table 1b:** Demographics for chronic obstructive pulmonary disease cases, from unrelated individuals of European ancestry in UK Biobank derived using logistic regression analyses. (Odds ratios and p values are adjusted for age and sex, as above).

| Demographic | COPD | Controls | OR | 95% CI |  | P |
| --- | --- | --- | --- | --- | --- | --- |
| N = 378,355 | 11,413 | 366,942 |  |  |  |  |
| Mean age at baseline (SD) | 61.9 (6.2) | 57.1 (8.0) | 1.10 | 1.09 | 1.10 | $<1 \times 10^{-15}$ |
| Mean age at diagnosis (SD) | 65.3 (7.4) |  |  |  |  |  |
| Male sex, N (%) | 6,245 (54.7%) | 167,910 (45.8%) | 1.38 | 1.32 | 1.43 | $<1 \times 10^{-15}$ |
| Townsend Deprivation Index (SD) | 0.21 (3.6) | -1.53 (2.95) | 1.19 | 1.19 | 1.20 | $<1 \times 10^{-15}$ |
| Pollution (SD) | 28.0 (7.7) | 26.2 (7.4) | 1.04 | 1.03 | 1.04 | $<1 \times 10^{-15}$ |
| Smoking status |  |  |  |  |  |  |
| Never smoker | 1,806 (15.8%) | 201,987 (55.1%) |  |  |  |  |
| Former smoker | 5,480 (48.0%) | 128,242 (35.0%) | 3.96 | 3.75 | 4.18 | $<1 \times 10^{-15}$ |
| Current smoker | 3,719 (32.6%) | 32,022 (8.73%) | 15.0 | 14.1 | 15.9 | $<1 \times 10^{-15}$ |
| Missing | 408 (3.6%) | 5,104 (1.3%) |  |  |  |  |
| Income (SD) | 1.77 (0.97) | 2.66 (1.19) | 0.53 | 0.52 | 0.54 | $<1 \times 10^{-15}$ |
| Mean FEV (SD) | 2.01 (0.72) | 2.77 (0.77) | 0.15 | 0.14 | 0.15 | $<1 \times 10^{-15}$ |
| FEV1 percent predicted (SD) | 71.2 (23.7) | 91.1 (22.6) | 0.96 | 0.96 | 0.96 | $<1 \times 10^{-15}$ |
| Mean FVC (SD) | 3.04 (0.93) | 3.66 (1.00) | 0.34 | 0.33 | 0.35 | $<1 \times 10^{-15}$ |
| FVC percent predicted (SD) | 107.6 (30.4) | 120.2 (29.6) | 0.98 | 0.98 | 0.98 | $<1 \times 10^{-15}$ |
| Physical activity score (SD) | 7.12 (1.31) | 7.41 (1.13) | 0.80 | 0.79 | 0.81 | $<1 \times 10^{-15}$ |
| Participants deceased, N (%) | 1,877 (16.5%) | 10,977 (3.0%) | 4.59 | 4.35 | 4.85 | $<1 \times 10^{-15}$ |

The extracted genetic variants, recoded to 0, 1 or 2 according to the number of telomere length associated alleles, were used to create a genetic risk score (GRS) for telomere length for each individual. The variants were weighted by their effect size (ß-coefficient) obtained from the primary GWAS (Equation 1), where each beta value reflects the standard deviation change (1-SD) in leukocyte telomere length per copy of the effect allele. The weighted score was then rescaled to reflect the number of trait raising alleles available (Equation 2).

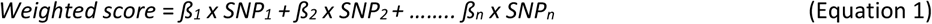

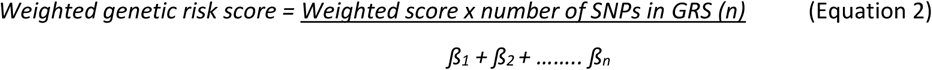

### Mendelian Randomisation

Mendelian randomisation (MR) was used to investigate causality between telomere length and incidence of IPF and COPD. MR relies on several general assumptions [23] which are applied in this case as follows:

a. the telomere length genetic variants are robustly associated with absolute leukocyte telomere length
b. the telomere length genetic variants are not associated, independently of their effects on telomere length, with confounding variables for IPF or COPD
c. the telomere length genetic variants are only associated with IPF or COPD via their effect on telomere length

In this study, we employed several MR approaches. First, we investigated the association between IPF or COPD and the telomere length genetic risk scores in the unrelated data set of 379,708 individuals using logistic regression models. Ancestral principal components (as previously described [24]) were included as covariates in the analysis to control for residual population structure and we also adjusted for baseline age, sex and UK Biobank assessment centre.

We next performed two-sample MR using the BOLT-LMM algorithm for mixed model association testing. The seven genetic variants associated with telomere length were extracted. A standard Inverse Variance Weighted (IVW) instrumental variable analysis was performed, along with two methods that are more resistant to pleiotropy: MR-Egger [25] and Median MR [26]. The IVW method regresses the effect sizes of variant outcome associations (here telomere length associated variants vs incidence of IPF or COPD) against effect sizes of the variant risk factor associations (here telomere length associated variants vs telomere length). Variant risk factor associations were taken from the primary GWAS of telomere length [14]. If no heterogeneity is detected amongst the causal estimates, the IVW analysis is carried out under a fixed effect model with the assumption of no horizontal pleiotropy. Alternatively, if heterogeneity is found amongst the causal estimates, a random effects model is implemented and the approach assumes that:

a. Either the strength of the association of the genetic instruments with the risk factor is not correlated with the magnitude of the pleiotropic effects
b. Or the pleiotropic effects have an average value of zero

In contrast, the MR-Egger method uses a weighted regression with an unconstrained intercept to remove the assumption that all genetic variants are valid instrumental variables and is therefore less susceptible to confounding from potentially pleiotropic variants that have a stronger effect on the incidence of IPF compared to their effect on telomere length. The Median-MR method is also more resistant to pleiotropy; it takes the median instrumental variable from all variants included and is therefore robust when <50% of the genetic variants are invalid. Given these different assumptions, if all methods are broadly consistent this strengthens the causal inference. Details of the R code for the 2-sample IVW, MR-Egger and Median-MR analyses are available in Bowden *et al* [26, 27].

Two sample MR in our IPF and COPD replication cohorts used summary GWAS statistics, the majority of which excluded UK Biobank data (77% for IPF and 100% FOR COPD).

Additional sensitivity analyses were performed to check the robustness of our results. First, the MR was repeated, excluding rs2736100 which has a reported link with IPF [28]. Second, we repeated the regression analyses in the unrelated subset of individuals using sex specific genetic risk scores (see Supplementary Material, Section 2). Third, we checked sensitivity of regression analyses for COPD to case description, since the definition of COPD cases in our replication cohort was based on lung function data (FVC% predicted <80%, FEV1/FVC <0.7) rather than reported diagnosis and recent evidence has shown that a spirometric definition identifies more cases [29]. Percent predicted spirometry values were generated in UK Biobank data using the Global Lung Initiative reference equations [30].

## Results

### Demographics

The demographics of the unrelated 1,133 IPF ‘narrow’ cases and 11,413 COPD cases are summarised in Table 1. Strong associations were noted between IPF and a range of demographic and environmental variables. Briefly, older age, male sex, lower socioeconomic position, ever smoking, reduced lung function and reduced exercise were associated with a higher odds ratio for IPF and also for COPD. Similar associations were noted for the 1,353 IPF ‘broad’ cases and also in the larger cohort of related individuals (Supplementary Material Section 3). Mortality figures were calculated with death as the outcome variable in each case.

### Telomere length polygenic genetic risk score

Using the 7-variant telomere length genetic risk score in our UK Biobank unrelated cohort, we demonstrated that the telomere length GRS was associated with higher odds of IPF in our ‘narrow’ definition group of 1,133 IPF cases; odds ratio (OR) = 1.11 [95%CI: 1.07-1.15], P = 3.1×10^−8^, Figure 2. Repeating this for the broader definition IPF, containing 1,353 cases, did not greatly alter this finding, OR = 1.09 [95%CI: 1.05-1.13], P = 4.7×10^−7^ (Table 3).

**Figure 2.**
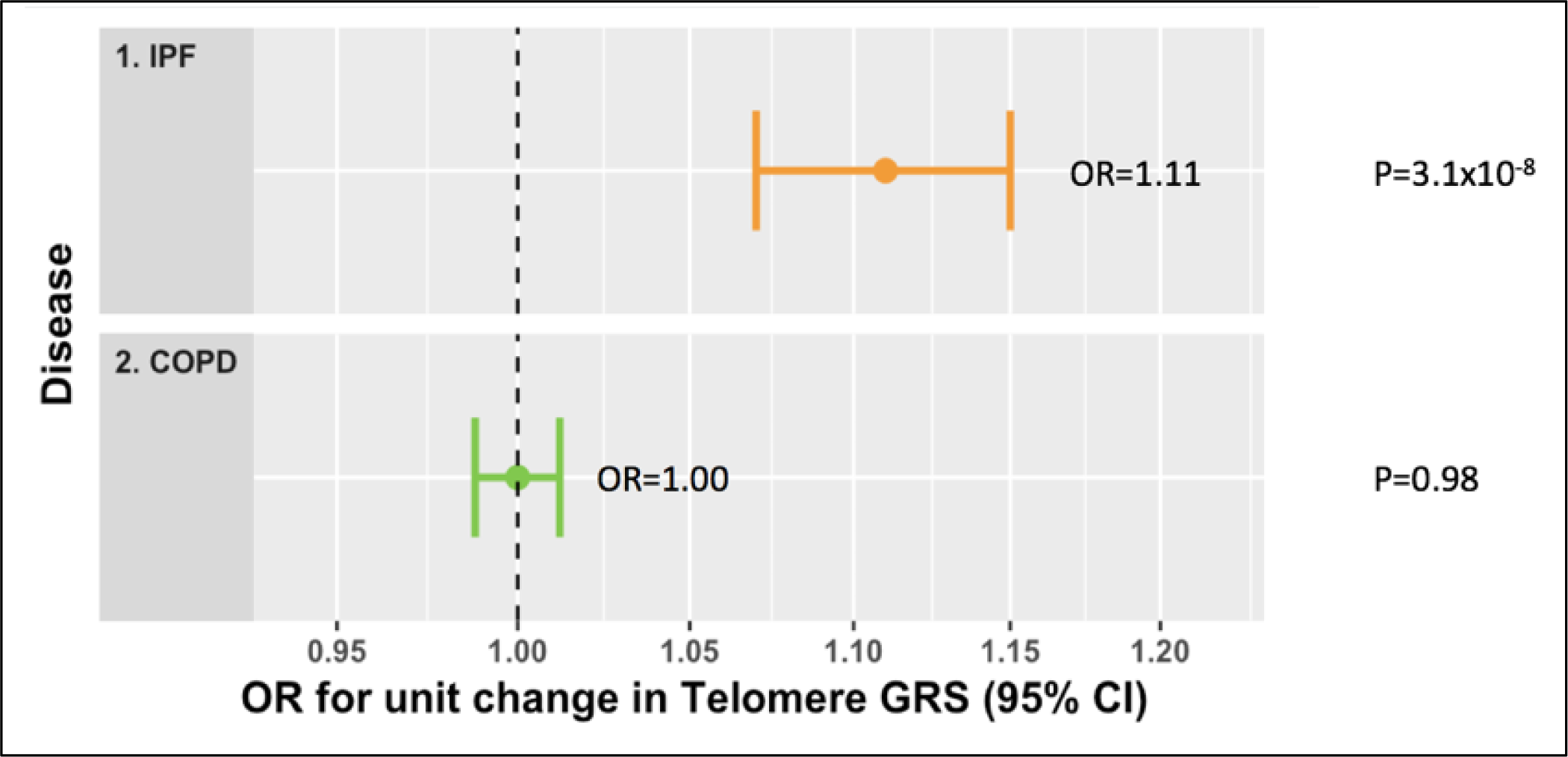
Logistic regression results showing significant disease risk odds ratio for IPF (‘narrow’) but not for COPD cases for a unit change in telomere length genetic risk score compared with controls in UK Biobank. An odds ratio that is significantly different to 1 indicates that odds of disease are influenced by telomere length.

In contrast, no association was seen for our COPD group of 11,413 cases (OR 1.000 [95%CI: 0.99-1.01], P=0.98), Figure 2.

With the 6 variant GRS (excluding rs2736100), the results were slightly attenuated but the GRS remained associated with IPF; for the ‘narrow’ group OR = 1.08 [95%CI: 1.04-1.13], P = 2.3×10^−4^ and for the broader definition IPF group OR = 1.06 [95%CI: 1.02-1.10], P = 2.5×10^−3^.

Due to the strong association between IPF incidence and male sex (OR = 1.94 [95%CI: 1.71-2.19], P <1×10^−15^; Table 1a), we used separate genetic risk scores for each sex, created from the 7 SNP sex specific beta values for telomere length. Repeating the MR generated similar results for males and females (Table 2).

**Table 2:** Evidence from UK Biobank data suggesting a causal role for telomere length in IPF but not in COPD. Associations between shorter telomere length genetic risk scores and disease incidence in IPF (‘narrow’), IPF (‘broad’), IPF males & IPF females (using the sex adjusted genetic risk scores) and COPD for genetic instruments derived from 7 and 6 variants. Results were adjusted for registration age, principal ancestral components and assessment centre and also for sex in mixed sex groups.

| Case Group | Genetic instrument | N cases (controls) | Odds ratios (95%CI) for IPF vs control per weighted allele of the GRS | P |
| --- | --- | --- | --- | --- |
| IPF 'narrow' All | 7 SNP | 1,133 (366,942) | 1.11 (1.07,1.15) | 2.1x10<sup>-8</sup> |
| IPF 'narrow' All | 6 SNP - no rs2736100 | 1,133 (366,942) | 1.08 (1.04, 1.13) | 2.5x10<sup>-4</sup> |
| IPF 'broad' All | 7 SNP | 1,353 (366,942) | 1.09 (1.05, 1.13) | 4.9x10<sup>-7</sup> |
| IPF 'broad' All | 6 SNP - no rs2736100 | 1,353 (366,942) | 1.06 (1.02, 1.10) | 2.7x10<sup>-3</sup> |
| IPF 'narrow' Men only | 7 SNP male grs | 718 (174,254) | 1.10 (1.05, 1.15) | 2.4x10<sup>-5</sup> |
| IPF 'narrow' Women only | 7 SNP female grs | 415 (204,321) | 1.12 (1.05, 1.19) | 2.2x10<sup>-4</sup> |
| COPD (our definition) | 7 SNP | 11,413 (366,942) | 1.000 (0.988, 1.012) | 0.98 |
| COPD (our definition) | 6 SNP | 11,413 (366,942) | 1.002 (0.989, 1.016) | 0.71 |
| COPD (lung function definition) | 7 SNP | 11,413 (366,942) | 1.003 (0.995, 1.010) | 0.48 |

In our sensitivity studies, we repeated the analysis for COPD using a lung function definition and the results were very similar (Table 2).

### Two-sample MR analysis

Two sample MR provided evidence that shorter telomere length causes IPF. In the UK Biobank, a genetically instrumented one standard deviation shorter telomere length was associated with higher odds of IPF; using the IVW method, in the narrow IPF group, OR = 4.19 [95% CI 2.26-7.77], P = 0.0031 and in the broad IPF group OR = 3.28 [95% CI 1.77-6.08], P = 0.0093. There was no evidence of a causal relationship in COPD (OR = 1.07 [95% CI 0.88-1.30], P = 0.51), (Figures 3 & 4 and Table 4, Supplementary Material). For the IPF groups, all methods were directionally consistent and the Egger method provided no evidence of pleiotropy (Egger intercept P-value: 0.45 in IPF ‘narrow’ and 0.47 in IPF ‘broad’).

**Figure 3.**
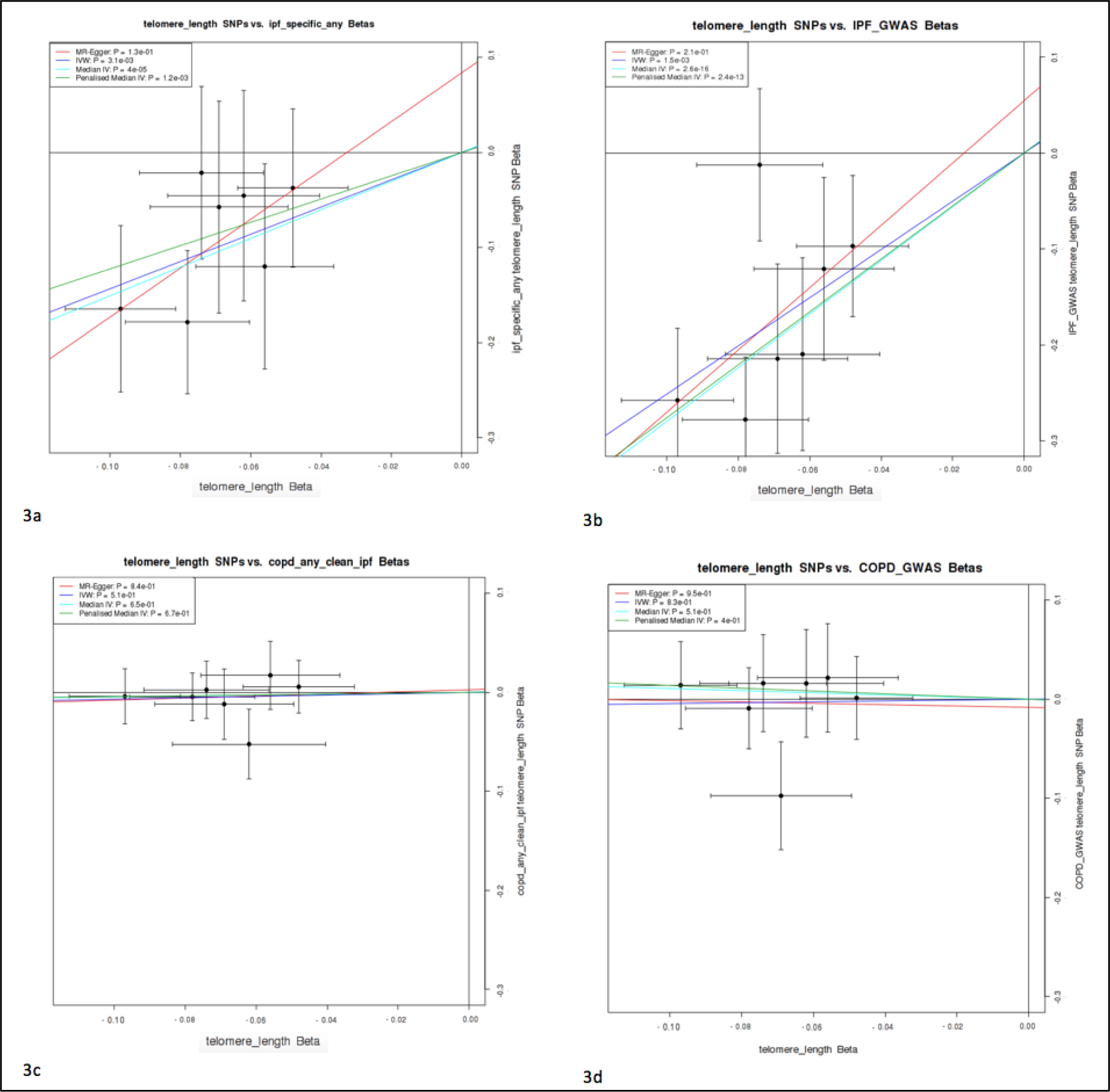
Two-sample MR results for IPF and COPD showing evidence of telomere length causality in IPF but not in COPD. Graphs show the strength of the relationship between disease incidence and telomere length SNPs on the y axis against the telomere length association from previous GWAS for each SNP on the x axis. A non-zero gradient to the lines, with significant p values shown in the top left-hand box for the different MR models used, is evidence of causality of telomere length for disease. Results shown are for all seven telomere variants for (a) IPF (‘narrow’) in UK Biobank, (b) IPF Replication Cohort, (c) COPD in UK Biobank, (d) COPD Replication Cohort

### MR results from replication cohorts

To provide further evidence for or against the causal role of telomere length in IPF, we used the summary statistics from the replication cohort data from 2,668 IPF patients and 8,591 controls. Again, two-sample MR provided evidence of a causal role for shortened telomeres in IPF. A 1-SD decrease in telomere length was associated with OR = 12.3 [95% CI: 5.05-30.1], P=0.015 (Figures 3 & 4 & Table 3). Similar associations were noted using the MR methods that are more robust to pleiotropy, and the Egger intercept provided no evidence of pleiotropy (P-value: 0.75).

Results from the COPD replication cohort provided no evidence of a causal relationship in COPD ((OR = 1.04 [95% CI 0.71-1.53], P = 0.83), (Figures 3 & 4 and Table 3).

### Meta-analysis of UK Biobank and replication cohort data

Meta-analysis of the IVW model estimates for IPF from the UK Biobank and the replication cohort suggested a causal relationship with a 1-SD shorter telomere length leading to OR= 5.81 [95% CI: 3.55-9.50], P= 2.19×10^−12^ for IPF. Using this model there is some evidence of heterogeneity between cohorts (heterogeneity P= 0.05). In comparison, meta-analysis of IVW estimates for COPD from the UK Biobank and the COPD replication cohort provided no evidence of any causal relationship for telomere length, with OR= 1.07 [95%CI:0.90-1.27, P=0.46, heterogeneity P=0.91]. Independent cohort and combined meta-analysis results are shown in Figure 4.

**Figure 4.**
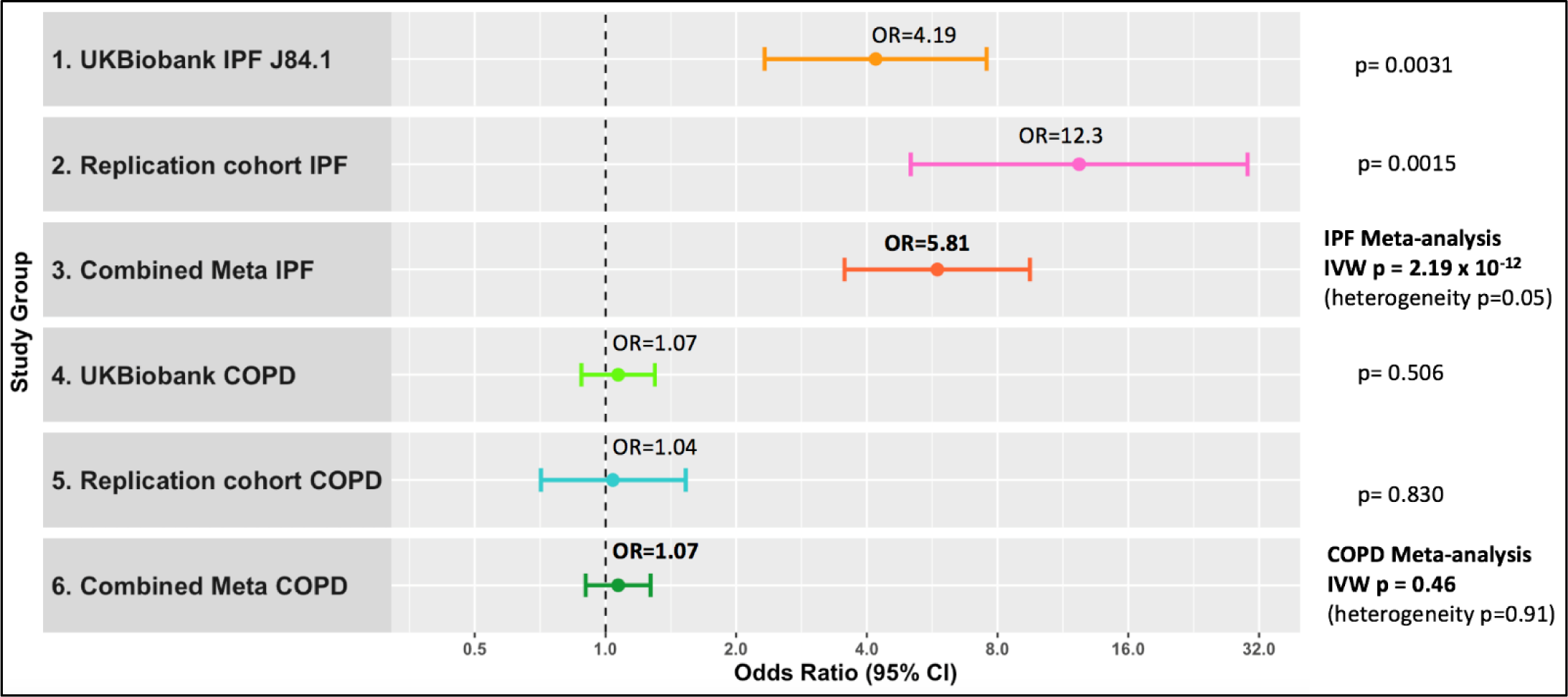
Meta-analysis results for IPF and COPD in UK Biobank and replication cohorts showing significant evidence of telomere length causality in IPF and not COPD across cohorts. Odds ratios and 95% confidence intervals for IPF (‘narrow’) in UKB, IPF Replication Cohort, IPF meta-analysis, COPD in UK Biobank, COPD Replication Cohort and COPD meta-analysis using IVW Method.

Similarly, meta-analysis results for MR-Egger estimates for IPF suggested a causal relationship for telomeres in IPF with a 1-SD shorter telomere length leading to OR= 15.8 [95% CI: 1.48-169], P= 0.02, with heterogeneity P= 0.80. Again, meta-analysis results using MR-Egger estimates for COPD provided no evidence of any causal relationship for telomere length, with OR= 1.07 [95%CI:0.45-2.56, P=0.88, heterogeneity P=0.87]. independent cohort and combined meta-analysis results are shown in Figure 6 of the supplement.

## Discussion

Using a Mendelian randomisation approach, we have shown that decreased telomere length is associated with increased risk of IPF but not COPD. The majority of our findings persisted when they were derived using models that made allowance for violations of MR assumptions, such as confounding by pleiotropy, with outcomes which were broadly consistent. These data therefore provide robust evidence of a causal link from short telomeres for idiopathic pulmonary fibrosis, while suggesting divergent underlying disease mechanisms in COPD.

Both IPF and COPD are exemplars of age-related disease, with increasing focus on the impact of accelerated ageing and particularly the pathogenic role of cellular senescence. This is a complex cellular programme characterised by cell cycle arrest and morphological/phenotypic changes, and can be broadly subdivided into replicative and cellular senescence [31]; the former results from intrinsic cellular events, including telomere shortening, while the latter can be driven by various stimuli including oxidative stress and DNA damage [32]. Increasing senescent cell burden has been attributed to both IPF and COPD, with evidence from several cellular compartments including type II alveolar epithelium and fibroblasts (see [33] for a comprehensive review). Both IPF and COPD have significantly shorter leukocyte telomere length [5, 6]; shortened telomeres have also been identified directly within lung tissue from IPF patients [8, 34] but not in COPD [35]. In light of our findings, these observations suggest that telomere shortening can act as an intrinsic and systemic driving force of cellular senescence in IPF, akin to the relationship between telomere-associated driver mutations in familial PF. In contrast, extrinsic stimuli (such as cigarette smoke exposure) may have a greater influence on senescent cell burden in COPD. In both diseases, short leukocyte telomere lengths could also result from immunosenescence, or possibly hypoxemia, given the reported association between telomere length and PaO2 [36]. Notably, LTL in COPD does not appear to be influenced by smoking status (current vs ex-smokers [37]), although smoking *per se* can cause telomere attrition [38]. Of interest, a recent study by Kachuri *et al* [39] used MR to investigate a causal role for longer telomeres in lung cancer, demonstrating an increased odds ratio for lung adenocarcinoma but not squamous cell carcinoma (SCC) or head and neck cancer. IPF is linked with an increased risk of lung cancer [40], but notably SCC rather than adenocarcinoma [41].

Evidence of a causal role for premature telomere attrition in IPF presents several significant opportunities. Approaches aimed at addressing inadequate telomere maintenance either universally or in the cells contributing most to the disease pathogenesis, may offer a therapeutic option. Restoration of telomere length is not as straightforward as simply upregulating telomerase, the enzyme that promotes telomere elongation, since this would promote the risk of cancer (already higher than average in IPF patients [40]), although clinical trials are underway using this approach in other conditions (see NCT04110964 at clinicaltrials.gov). In light of this, safe telomerase activation therapy is being explored in other medical fields such as cardiology, using transient delivery which avoids creating an environment in which increased telomerase persists. In a study by Ramunas *et al* reported early in 2015 [42], *in vitro* delivery of a TERT-encoding mRNA, avoiding the risk of insertional mutagenesis, to human fetal lung fibroblasts and myoblasts, successfully resulted in telomere extension and an increase in proliferative capacity.

Targeted delivery *in vivo* presents a very significant challenge to utilising such an approach in PF, but future developments may render this feasible. Diminution of the senescent cell burden in IPF is also currently being explored using senolytic drugs such as the combination of dasatinib plus quercetin, for which an open label Phase I Study has recently been reported [43]. Alternative approaches could include exploration of androgen therapy: testosterone has been used effectively to treat the telomere linked disease, aplastic anaemia, for many years and Danazol has shown some promise in the treatment of PF, with an apparent arrest in lung function decline over 36 months [44]. Androgens can also restore telomerase to normal levels in cells from telomere disease patients who are heterozygous for TERT gene mutations [45].

There are also existing well-known therapies which have a positive effect on promoting telomere length and which could be readily adopted as an important part of clinical practice for treating IPF patients. These include exercise [46], reduction of life stress [47] and mindfulness [48, 49]. Our data show that participants with IPF tended to have reduced physical activity (Table 1a), probably exaggerated by poor physical health, and we propose that carefully increased exercise could offer multiple benefits in terms of promoting chromosomal telomere length and also boosting fitness and mental health. Similarly, stress arising from difficult life circumstances such as social deprivation (which our data show is associated with IPF, Table 1a) can be relieved with practices such as mindfulness [48, 49]. Relatively simply practices which improve patient wellbeing may also have a fundamentally positive impact on reducing telomere attrition.

In summary, we have found evidence, in both our UK Biobank and replication cohorts, of a causal link between telomere length and sporadic IPF. We found no evidence of any link to COPD, a similar age-related disease, in either the UK Biobank or our COPD replication cohort. The identification of a cause behind human IPF leads us to new insights towards beneficial therapies for patients and routes to potential new treatments - which may lead us closer to preventing the disease in those with prematurely shortened telomeres, and ultimately providing a direction in our search for a cure.

## Data Availability

UK Biobank is an open access resource. The Resource is open to bona fide scientists, undertaking health-related research that is in the public good. Approved scientists from the UK and overseas and from academia, government, charity and commercial companies can apply to use the Resource.
Replication cohort summary statistics for the IPF GWAS are available via application at the URL below.
Replication cohort summary statistics for the COPD GWAS are available on request at the URL below.

https://www.ukbiobank.ac.uk/

https://github.com/genomicsITER/PFgenetics

https://www.ncbi.nlm.nih.gov/pmc/articles/PMC5381275/

## Acknowledgements and funding

A. Duckworth is funded by the GW4 MRC Doctoral Training Partnership.

M. Gibbons has received support to attend conferences and professional fees from Roche and Boehringer-Ingelheim

R. Allen is an Action for Pulmonary Fibrosis Research Fellow.

L. Wain holds a GSK/British Lung Foundation Chair in Respiratory Research.

J. Tyrrell is supported by an Academy of Medical Sciences (AMS) Springboard award, which is supported by the AMS, the Wellcome Trust, GCRF, the Government Department of Business, Energy and Industrial strategy, the British Heart Foundation and Diabetes UK [SBF004\1079].

This research has been conducted using the UK Biobank Resource (applications 9072 and 44046).

The authors would like to acknowledge the use of the University of Exeter High-Performance Computing (HPC) facility in carrying out this work.

The Research was partially supported by the National Institute for Health Research (NIHR) Leicester Biomedical Research Centre; the views expressed are those of the author(s) and not necessarily those of the National Health Service (NHS), the NIHR or the Department of Health

